# Cardiovascular disease risk and Lung cancer screening for Early Assessment of Risk (project CLEAR) in Missouri: A protocol for a mixed methods study

**DOI:** 10.1101/2025.05.21.25328014

**Authors:** Beryne Odeny, Isaac Che Ngang, Vaishnavi Mamillapalle, Mariam Balogun, Benjamin Bowe, Meera Muthukrishnan, Akila Anandarajah, Feng Gao, Aimee James, Graham Colditz

## Abstract

**Background:** Lung cancer and atherosclerotic cardiovascular disease (ASCVD) are leading causes of mortality in the United States, sharing common risk factors like smoking and age. Preventive care for these conditions is often siloed, leading to missed opportunities to prevent ASCVD-related mortality in lung cancer screening (LCS) patients. In various trial settings, patients undergoing LCS were more likely to die from ASCVD than from cancer; however, less than half of eligible patients got statin prescription. This study aims to understand the degree to which ASCVD risk assessment and prevention is done in routine “real-world” clinical settings providing LCS in Missouri. Our objectives are to determine the prevalence of statin eligibility, statin prescription and explore factors, disparities and barriers to statin therapy in people undergoing LCS.

**Methods:** A parallel convergent mixed-methods design will be used. Quantitative data from people undergoing LCS at Barnes-Jewish Healthcare System will be extracted from Epic electronic medical records (EMR) from January 2022-December 2023, capturing demographics, socioeconomic factors, and clinical data including ASCVD risk factors and statin use. We anticipate gathering about records for approximately 8,000 patients. Qualitative data will be gathered from in-depth interviews with up to 15 healthcare providers to identify barriers, facilitators and recommendations for enhancing statin therapy in people undergoing LCS.

**Discussion:** The risk of mortality is drastically heightened in people undergoing LCS who are not receiving statin therapy, and thus there is an urgent need to identify those who are eligible but not receiving ASCVD prevention. Promoting ASCVD risk assessment and prevention in LCS programs has the potential to mitigate the dual burden of ASCVD and lung cancer in high-risk populations who smoke cigarettes. This study will demonstrate the extent of the gap in statin therapy among LCS and provide insights into disparities, barriers and recommendations for integrating ASCVD risk assessment and prevention with LCS.

**Conclusion:** This study will provide real-world baseline data on statin therapy among people undergoing LCS in Missouri, and will highlight disparities, barriers and recommendations for integrating ASCVD prevention in LCS.

## Introduction

Lung cancer and atherosclerotic cardiovascular disease (ASCVD) remain two of the leading causes of death both in the United States and globally. Lung cancer accounts for about 25% of all cancer deaths, while cardiovascular diseases, including ASCVD like myocardial infarction and stroke, are the primary cause of mortality in older adults, particularly those with risk factors such as smoking, hypertension, and obesity [1-5]. Preventive measures such as lung cancer screening (LCS) using low dose computed tomography (CT) and ASCVD risk assessment and prevention with statin therapy (a drug used to reduce the risk of ASCVD) have significantly reduced mortality from these conditions [6,7]. However, cardiovascular events remain the leading causes of deaths among people undergoing LCS [8-10]. LCS can show any findings that could then lead to reconsidering statin therapy: for example, the low-dose CT may show moderate to major coronary calcification which can be a trigger for further conversation around cardiac health. The fact that LCS services and ASCVD risk assessments are typically siloed in different clinical settings and often provided by different providers may contribute to missed opportunities for delivering ASCVD prevention to people undergoing LCS [9].

Clinical trials and studies from the United Kingdom (UK) and the United States (US) have revealed that a significant proportion of people undergoing LCS who are eligible for statins have never received stain therapy [10-15]. A UK-based study showed that more than half of statin-eligible individuals in a LCS cohort were not on statin therapy despite their elevated ASCVD risk [11]. Similarly, a study in North Carolina found that approximately 60% of statin-eligible patients undergoing LCS were not prescribed statins [12]. These findings highlight a critical gap in ASCVD prevention despite high ASCVD mortality rates in people undergoing LCS. We aim to bridge this gap by examining the prevalence of statin eligibility and prescription among patients undergoing LCS in a routine “real-world” clinical settings.

Information on ASCVD risk assessment and prevention in LCS contexts in Missouri is lacking. Given the high prevalence of smoking [16] and ASCVD [17] in the region, it is essential to evaluate the current state of ASCVD risk assessment and prevention with statin therapy for this high-risk population. To address this dearth of knowledge, we propose a parallel convergent mixed methods study to determine the baseline prevalence of statin-eligible individuals in LCS programs and the proportion of statin-eligible individuals missing statin therapy in Missouri. We will investigate covariates and disparities associated with statin therapy in LCS services. We will qualitatively explore perceived barriers, facilitators, and recommendations for promoting ASCVD risk assessment and prevention with statin therapy in LCS services in Missouri. This study will highlight any gaps in statin prescription among patients accessing LCS and recommend strategies for ensuring that all patients have CVD risk assessment and statin therapy, if eligible. We hypothesize that statin prescription among people referred for LCS will be suboptimal, and disparities across race and ethnicity, socioeconomic status, and geographic regions (i.e., rural versus urban) will exist.

### Conceptual framework

Our research maps on the **“Exploration**” phase of the EPIS (Exploration-Preparation-Implementation-Sustainment) framework [18] which involves assessing the current gaps in ASCVD risk assessments among people undergoing LCS and identify disparities in preventive care. This phase will involve engaging health providers to understand the barriers and facilitators to statin prescription among people undergoing LCS in Missouri. Within this exploration phase, we will apply the domains of the Socio-ecological Model (SEM) to guide inquiry into, and analysis of, healthcare provider perspectives. The SEM is a broad framework that facilitates the understanding of multi-level factors that affect health outcomes including intrapersonal (individual), interpersonal, organizational, public policy, and community factors [19]. Figure 1 demonstrates how we integrate the EPIS implementation framework and SEM. Future research will focus on the remaining phases of the EPIS framework, from “preparation” to “sustainment.”

**Figure 1.**
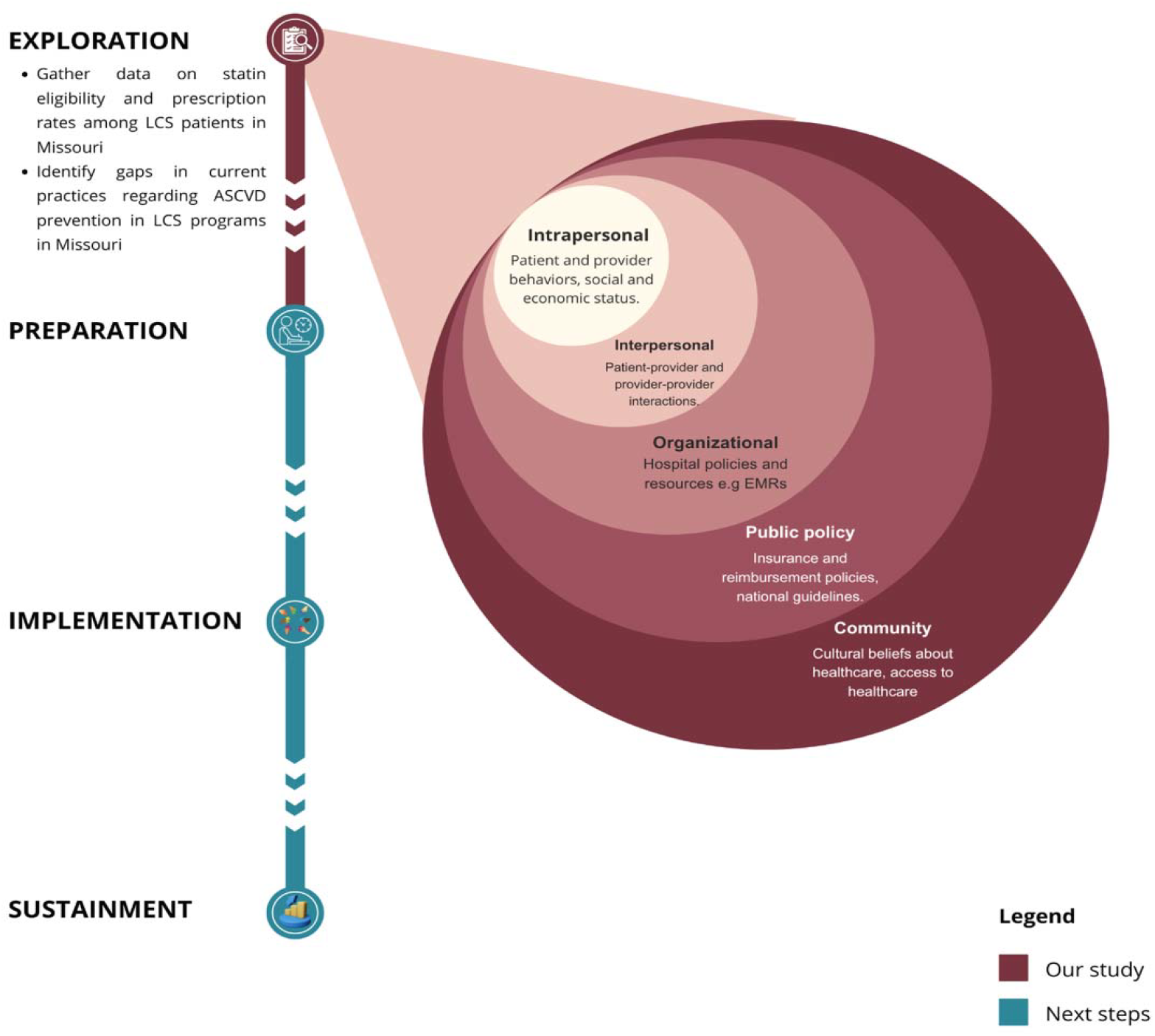
Conceptual framework of the study- EPIS and SEM frameworks

## Methods

### Study design

We use a parallel convergent mixed-methods study design, that triangulates findings from quantitative and qualitative data. The study will be conducted at the Barnes-Jewish Healthcare System (BJC) which operates in Missouri and Illinois. Both lung cancer screening and ASCVD prevention care are offered in BJC albeit at different departments. To be eligible for this study, participants must receive primary care at BJC. The study consists of two parts: (1) a cross-sectional study, in which electronic health data will be obtained through extraction from Epic, which is the electronic medical record (EMR) platform used in BJC, and (2) qualitative healthcare provider in-depth interviews through which we will gather insights, experiences and perspectives of healthcare providers (Figure 2).

**Figure 2.**
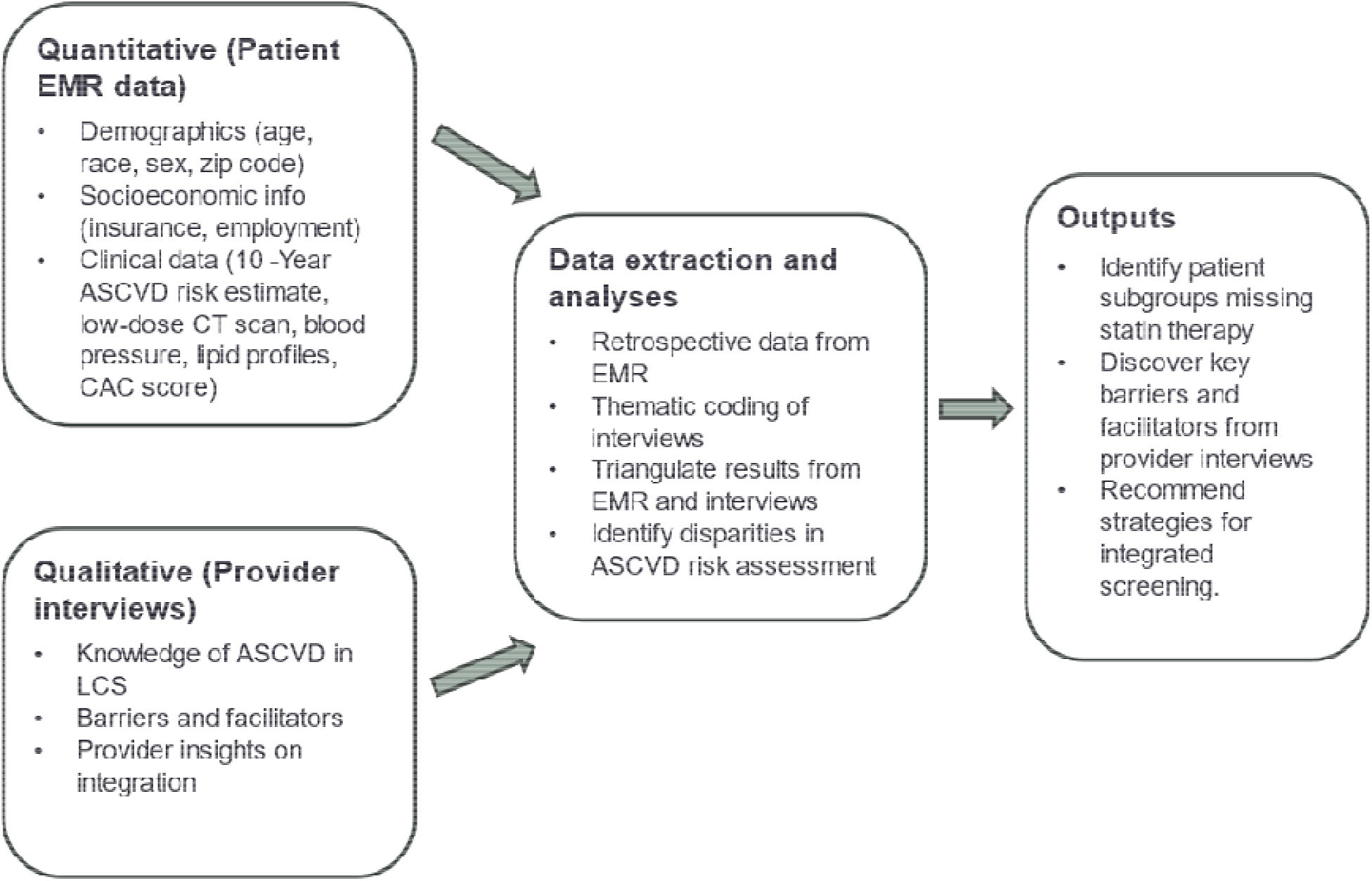
Overview of the study

### Study objectives

1. To determine the baseline prevalence of statin-eligible individuals and the proportion of statin-eligible individuals who are missing statin therapy in LCS program in Missouri.
2. To assess factors associated with statin therapy and whether disparities exist in statin prescription among statin-eligible individuals receiving LCS services in Missouri.
3. To explore barriers, facilitators and recommendations for promoting ASCVD risk assessment and prevention with statin therapy in LCS services in Missouri.

### Study Procedures

#### Cross-sectional study of Epic data

Quantitative data will be obtained from Epic for those patients who have accessed LCS (eligibility criteria is provided in Section 3.1) i.e., individuals between the ages of 50 and 80, who have a 20 pack-year smoking history, and currently smoke or quit within the past 15 years. All individual-level data that streams in from different departments is recorded in Epic and linked back to the patient using a unique medical records number (MRN).

We will extract demographic data, socioeconomic information (such as employment, insurance type,), geographic location (rural vs. urban based on zip code of residence data if available), race and ethnicity, smoking history, medications, clinical history including procedures like low-dose CT scan, blood pressure, lipid profiles, glucose levels, and ASCVD risk enhancers such as family history and metabolic syndrome. The dates of any statin prescribed over the last six years, including information on whether it was prescribed prior and/or after to a patient undergoing LCS will be collected using the dates of procedures and prescriptions. We will identify any new prescriptions, including those administered this year in 2024. Information on whether patients were referred to their primary care provider and whether specialist care was accessed (e.g. pulmonologist) will be extracted. Using zip code data, we will determine the multiple deprivation index (MDI) [20] associated with individual’s residence as a marker of socioeconomic status and access to healthcare. Data on race and ethnicity typically includes a field for recording the patient’s self-reported race and ethnicity. The racial and ethnic data we anticipate gathering include American Indian or Alaska Native, Asian, Black or African American, White and other Pacific Islander. The information on race and ethnicity will be used to examine potential disparities in cardiovascular risk assessment and prevention therapy across different racial groups. For geographical location, we will use zip code data to determine rural-urban residence.

If available, radiological findings on the low-dose CT scan, including the coronary artery calcification (CAC) score will be captured. If available, EMR-computed 10-year ASCVD risk estimate will also be captured; it is a composite score based on the risk factors (calculated using the Pooled Cohort Equation [21-23].

For the 10-year ASCVD risk estimate, we acknowledge that these estimates may not always be pre-calculated and stored within the EMR. To address this, we will employ a two-pronged approach:

- We will prioritize collecting existing, pre-calculated risk estimates whenever available.
- If the pre-calculated estimate is unavailable, we will utilize the readily available data in the EMR – age, sex, race and ethnicity, cholesterol, blood pressure, smoking and diabetes status – to calculate the risk estimates manually using the Pooled Cohort Equation (PCE) tool [21-23].

The CAC score availability is not essential for achieving aim 1 or 2. However, including it in the analysis, if available, would provide valuable additional data for further corroboration with the 10-year ASCVD risk estimates.

There are 4 ASCVD risk categories: low, borderline, intermediate or high. Statin prevention is strongly recommended (class I recommendation) for high and intermediate risk groups and is favorably considered (class IIa recommendation) for those with borderline risk plus ASCVD risk enhancers such as family history of premature ASCVD, certain inflammatory conditions, and ethnicity [21-22]. For statin prescription before ASCVD diagnosis, we will differentiate between statin use for prevention versus treatment using diagnostic codes using the International Classification of Diseases, Tenth Revision, Clinical Modification (ICD-10) [24] or Current Procedural Terminology (CPT) codes [25].

#### Qualitative in-depth interview (data collection)

We will conduct up to 15 in-depth interviews with healthcare providers who oversee, offer, or refer individuals to LCS (including primary care physicians, nurses, advanced practice providers and specialists like pulmonologists). To enhance the richness and generalizability of the qualitative data, we plan to recruit providers of varying specialties at BJC. We will prioritize recruiting providers from one main location - the main BJC hospital. Providers’ experience does not have to be confined to the specific timeframe of the quantitative data analysis. The qualitative interviews aim to understand provider experiences and perspectives within the health system, not necessarily specific to the timeframe of the EMR data. Before beginning the interview, verbal consent will be obtained. Virtual 30-minute Zoom or in-person interviews will be conducted based on provider preference. Interviews will address clinical workflows for LCS and CVD assessments, suggestions for integrating these workflows, and any potential barriers and facilitators (refer to S1 file for the interview guide). Data will be collected until data saturation is achieved. We will determine that data saturation has been reached when no new significant themes or insights are identified in subsequent interviews, suggesting that we have captured the breadth of relevant experiences and perspectives. Interviews will be audio-recorded, professionally transcribed, and de-identified before review.

#### Ethical Approval

We obtained ethical approval from the Washington University Institutional Review Board. We received a waiver of consent for electronic record data extraction and were have been approved to obtain verbal consent from health providers.

### Quantitative data analysis

#### Statistical analysis

We anticipate reviewing approximately 8,000 patient records. We will use descriptive statistics to summarize the distribution of patient characteristics. We will use proportions to summarize binary data such as whether a statin has been prescribed. We will calculate the proportion of statin-eligible individuals, the proportion of statin-eligible individuals receiving preventive statin prescription, and their 95% confidence intervals, calculated both in overall LCS accessing individuals and in subgroups stratified by racial and ethnic group, rural versus urban location, and socioeconomic status. We will use multivariable logistic regression models to estimate associations between variables and statin prescription.

We will adjust for relevant confounders such as age, sex, and insurance status. We will explore potential effect modifiers, such as race and ethnicity, that might influence the relationship between statin-eligibility and statin prescription. To assess disparities in statin prescription, multivariable logistic regression models will be used to assess associations between statin prescription and diverse groups (e.g., racial and ethnic groups) and to estimate the group-specific rates of statin prescription among statin-eligible individuals.

To handle missing data, we will conduct a thorough data cleaning process to identify and understand patterns of missing values. If missing data is considered randomly missing, we will consider using multiple imputation techniques to estimate missing values based on available data. All analyses will be conducted in statistical packages R and SAS.

#### Power analysis

This study will extract data from all individuals aged 50 to 80 years attending BJC that have accessed lung cancer screening services from January 2022-December 2023 and have ASCVD risk factor data in the EMR. A recent snapshot of lung cancer screening electronic medical data identified approximately 4,000 current or former smokers who completed LCS in 2023. Using N=5000 subjects as a conservative projection for total number of LCS accessing individuals during 2022-2023, the designed sample size will allow us a high precision to estimate the “true” prevalence of statin-eligible individuals and to identify associated demographic and clinical characteristics (**Objective 1**). For example, this allows us ≤0.7% measurement error to estimate the overall prevalence of statin-eligible individuals. For a risk factor with 10% prevalence (i.e., N=500 vs. N=4500), this provides 80% power at 2-sided alpha=0.05 to detect an effect size of 0.13 (in terms the difference of arcsine transformed proportions). Similarly, it provides 80% power at 2-sided alpha=0.05 to detect an effect size of 0.08 for a risk factor with 50% prevalence (i.e., N=2500 vs. N=2500). The projected sample size also provides a good precision to estimate the “true” proportion of statin-eligible patients accessing statin therapy and to assess its disparity across racial and SES subgroups (**Objective 2**). Assuming 43% prevalence (as supported by a study conducted at UK) of statin-eligible individuals in LCS, for example, N=∼2000 statin-eligible individuals provide ≤1.1% measurement error to estimate the “true” proportion of accessing statin therapy among statin-eligible individuals [11].

### Qualitative data analysis

We will conduct a thematic analysis employing mixed inductive and deductive approaches, grounded in the Socio-Ecological Model [19], i.e., this framework will guide the qualitative processes including data collection and analysis. This will help us organize and interpret the qualitative data within the context of the research question (Fig 1).

Inductive coding will be used to identify new emerging themes from the interview data. This allows us to discover new concepts and patterns that may not have been explicitly anticipated in the conceptual framework. Concurrently, we will use deductive coding, working with predetermined deduced codes based on existing literature and the conceptual framework, to guide our coding process and ensure structure in our analytical approach.

We will explore key barriers, facilitators, and recommended strategies for increasing appropriate prescription of statins. We will have at least 2 coders to enhance reliability and trustworthiness of the coding process. Coding discrepancies will be reconciled through discussion and consensus. Overarching themes will be explored and discussed. We will use NVivo v.14 qualitative data analysis software to facilitate the coding process and to manage data, codes and emerging themes.

#### Triangulating quantitative and qualitative data

We will analyze and interpret the quantitative and qualitative data separately, and then triangulate the qualitative and quantitative findings by directly comparing and contrasting statistical results with qualitative findings and validating quantitative findings with qualitative data. We will identify areas of convergence, when quantitative data (e.g., observed low statin prescription rates) aligns with themes from in-depth interviews (e.g., identified barriers to statin prescription). This will strengthen the overall credibility of the findings. We will explore areas of divergence as well. If quantitative data suggests a disparity, and in-depth interviews reveal different reasons than anticipated, this can prompt us to delve deeper into the reasons behind the observed disparities. We will present quantitative data and qualitative data separately and report in stages.

## Discussion

This study provides a significant opportunity to improve health outcomes in patients undergoing LCS by investigating the state of ASCVD risk assessment and prevention in LCS programs. Integrating ASCVD risk assessment into LCS programs has been endorsed by the American College of Cardiology, in recognition of the potential for this synergistic approach to significantly reduce cardiovascular disease-related mortality [15]. The benefits of this integration are clear: by addressing two major health risks simultaneously, healthcare providers can deliver more comprehensive, efficient care, particularly for high-risk populations who may not otherwise engage with cardiovascular screening services. Moreover, studies have suggested that this integration can be achieved at minimal additional cost, making it a cost-effective strategy for improving public health [6].

This study aims to bridge the gap between these two prevention paradigms—LCS and ASCVD prevention with statin therapy. The first objective of this study is to determine the baseline prevalence of statin-eligible individuals undergoing LCS, as well as the proportion of these individuals who are not receiving statin therapy. Existing research in trial settings highlights a significant gap in cardiovascular care for patients accessing LCS. Our study will quantify this gap in routine, real-world settings in Missouri, providing crucial baseline data that will inform future interventions aimed at improving ASCVD prevention and statin uptake among people undergoing LCS.

In addressing the second objective, this study will assess whether disparities exist in statin prescriptions among individuals accessing LCS services. Previous research has demonstrated that racial and socioeconomic disparities significantly affect access to preventive cardiovascular care. Studies have shown that minority populations, including Black and Hispanic individuals, are less likely to receive statin therapy compared to their White counterparts, even when eligible [26]. Similarly, individuals from lower socioeconomic backgrounds or rural areas often face barriers to receiving preventive care, including limited access to healthcare providers and medications. By analyzing demographic and socioeconomic data including multiple deprivation indexes, this study will help identify specific subgroups disproportionately affected by gaps in care.

Our third objective involves exploring the perceived barriers, facilitators and recommendations to integrating or promoting ASCVD risk assessment and prevention in LCS settings. Studies have identified several barriers, including time constraints, lack of coordinated care between specialists, and uncertainty regarding ASCVD guidelines in cancer screening settings [24]. By gathering firsthand perspectives from healthcare providers, we aim to determine the specific factors that hinder or facilitate the adoption of ASCVD risk assessment among people undergoing LCS in Missouri. This information will support the development of targeted strategies to integrate LCS and ASCVD prevention services.

One of the key strengths of our study is its use of a parallel convergent mixed-methods design, which allows for the triangulation of both quantitative and qualitative data. The BJC has a robust LCS program with high volumes of patients. As such, the analysis of expansive volumes of patient records of ∼8,000 will provide robust, objective data on statin eligibility and prescription patterns. On the other hand, the qualitative interviews with providers will offer a deeper understanding of the numbers and estimates we observe and potential barriers and facilitators. This exploratory research will provide a foundation upon which to develop and test strategies to increase ASCVD prevention with statin therapy in people undergoing LCS.

We anticipate a few limitations with this study. First, the cross-sectional design reduces our ability to make causal inferences with regards to determinants or drivers of statin therapy uptake among people undergoing LCS due to confounding factors and other biases. We will not be able to determine whether statin prescription was coincidental or directly caused because of the low-dose CT. We will control for potential confounding factors statistically by employing multivariable regression analysis accounting for confounders and effect modifiers. Second, the single-center focus of this study limits generalizability of findings to diverse settings; however, we have a huge patient volume which increases applicability of findings to similar settings. Third, we are concerned about missing data; we will address this using complete case analysis or multiple imputation and sensitivity analysis, as appropriate.

In conclusion, this study is poised to make a significant contribution to holistic patient maintenance in cancer screening by providing crucial data on the intersection of LCS and ASCVD risk assessment and prevention. The study’s findings will not only quantify the current gaps in statin prescriptions for people undergoing LCS but will also uncover the multi-level barriers that prevent healthcare providers from offering statins to people undergoing LCS. This research will pave the way for future efforts to develop and implement integrated screening protocols for lung cancer and ASCVD prevention with the goal of reducing mortality and promoting health among high-risk populations.

## Data Availability

All data produced in the present study are available upon reasonable request to the authors

## ACKNOWLEDGEMENTS

We would like to thank Dr. Justin Sadhu for providing overall guidance on the methodological approach for this protocol.

## APPENDIX

### Supporting information

#### S1 File. Qualitative interview questions

1. Briefly describe your clinical or research role as it relates to lung cancer screening. Are you a primary care provider or provider of lung cancer screening services?
2. If possible, please briefly describe the clinical workflow for lung cancer screening or referral for lung cancer screening.
  a. How are eligible patients identified for lung cancer screening?
  b. Are there any self-referrals for lung cancer screening?
3. What are the barriers to referral or access to lung cancer screening services?
  a. Probe: Please describe provider, patient, health system, and societal factors.
4. What factors facilitate referral or access to lung cancer screening services?
  a. Probe: Please describe provider, patient, health system, and societal factors.
5. How is cardiovascular disease risk assessment and prevention integrated into the care of the patients who require lung cancer screening services?
  a. Probe: If any, what is the dedicated process for assessing and managing cardiovascular disease risk in patients who need lung cancer screening?
6. How are patients’ cardiovascular risks determined and documented?
  a. Probe: Are there Epic prompts or flags for patients with higher cardiovascular disease risk like those needing lung cancer screening?
7. If applicable to your experience, please indicate which cardiovascular disease prevention interventions you typically prescribe for high-risk patients?
  a. How aggressive is this management for patients who are eligible for lung cancer screening, compared to those who are not?
8. Do you have any recommendations for ensuring that all patients who are referred for lung cancer screening have been assessed for cardiovascular disease risk and provided statin therapy, if eligible?
9. What are the potential barriers to integrating cardiovascular disease risk assessment and statin therapy into lung cancer screening workflows or services?
10. Are there any practices, policies, or resources that could facilitate the integration of CVD risk assessment and statin therapy within lung cancer screening services to ensure that this high-risk group of patients do not miss statin prevention therapy?

